# Key stakeholder perspectives on implementation of mHealth and NCD- related interventions in Kenyan Emergency Departments

**DOI:** 10.64898/2026.06.01.26354650

**Authors:** Grace Soma, Luz Mercado, Jessica Rayo, Mari Armstrong-Hough, Steven Bernstein, Lorien Abroms, Christine Ngaruiya

## Abstract

**Background:** Emergency Department (ED) populations are a high-risk group that are opportune for interventions targeting NCDs and NCD risk factors, like tobacco use. Mobile health (mHealth)interventions such as Text2Quit, a novel text message-based mHealth tool addressing tobacco cessation in the US, have demonstrated effectiveness for tobacco cessation and for ED-based mHealth interventions in High Income Countries (HIC).

To successfully adapt and implement such mHealth interventions in limited resource settings like African EDs, it is essential to examine the implementation climate and engage key stakeholders. These implementers provide invaluable insight to understand healthcare system level factors that affect adoption, implementation and maintenance of the interventions.

**Methods:** We conducted 12 semi-structured key informant interviews (KIIs) with ED administrators and staff including 2 departmental heads, 5 medical doctors, 3 nurses, and 2 clinical officers at a national referral hospital in Kenya. This was guided by RE-AIM framework indicators of “Adoption”, “Implementation”, and “Maintenance” (eg feasibility of intervention integration, and suggestions to improve implementation).

Interviews were conducted in English, recorded, professionally transcribed and translated, and analyzed using a constant comparative analysis approach, according to grounded theory principles.

**Findings:** Key informants were positive about the adoption of them Health intervention in Kenyan EDs and across different health facility levels in Kenya due to the perceived need for the program, facility and staff receptiveness and existing healthcare infrastructure to leverage. Recommended implementation strategies included follow-up mechanisms for program participants, inclusion of all healthcare cadres in implementation and increased sensitization and the use of champions. Barriers to Implementation in the ED included competing clinical priorities with emergency cases, limited staffing and shame associated with smoking.

**Conclusion:** Implementing a mobile health tobacco cessation program like Text2Quit is feasible and acceptable in Kenyan EDs when supported by targeted strategies.

## Introduction

Non-communicable diseases (NCDs) are a leading cause of mortality worldwide, accounting for approximately 75% of non-pandemic-related global deaths, with the majority occurring in low- and middle-income countries (LMICs) ^1^. Tobacco use remains a major preventable contributor to cardiovascular disease, chronic respiratory disease, stroke, cancer, and other NCDs, and kills more than 7 million people each year, including an estimated 1.6 million non-smokers exposed to secondhand smoke^1^. Although global tobacco consumption has declined substantially in many high-income countries due to comprehensive tobacco control policies, public health campaigns, and improved access to cessation services, tobacco use in Africa is projected to increase significantly over the coming decades^2^.

The WHO Framework Convention on Tobacco Control^3^(FCTC) and the MPOWER measures were developed to strengthen global tobacco control efforts through evidence-based policy and public health interventions^4^. The MPOWER package emphasizes six key strategies: monitoring tobacco use and prevention policies, protecting people from tobacco smoke, offering help to quit tobacco use, warning about the dangers of tobacco, enforcing bans on tobacco advertising and sponsorship, and raising taxes on tobacco products^4^. Despite progress in several tobacco control domains, implementation of tobacco cessation services remains uneven globally, particularly in LMICs where cessation infrastructure, provider training, and access to treatment remain limited^5^.

Emergency Department (ED) populations represent a particularly high-risk group for NCDs and related behavioral risk factors, including tobacco use. Tobacco use contributes substantially to conditions commonly encountered in ED settings, including cardiovascular disease, chronic obstructive pulmonary disease, asthma exacerbations, stroke, and tobacco-related malignancies. Prior studies conducted in Kenyan ED settings have demonstrated a disproportionately high burden of NCDs and associated risk factors compared to the general population, highlighting the ED as an important yet underutilized setting for preventive and behavioral health interventions^6^. Emergency medicine–based behavioral interventions, including Screening, Brief Intervention, and Referral to Treatment (SBIRT) models, have demonstrated the potential for ED encounters to serve as opportunities for preventive counseling and linkage to care among high-risk populations^7,8^.

Mobile health (mHealth) interventions offer a promising and scalable approach to tobacco cessation. Mobile phone–based smoking cessation interventions have demonstrated effectiveness in improving quit outcomes in randomized controlled trials^9^. Text2Quit, a bidirectional automated text message–based tobacco cessation intervention grounded in social cognitive theory, has demonstrated effectiveness in improving smoking cessation outcomes in the United States^10^. Although mHealth interventions have increasingly been applied within African contexts for communicable diseases such as HIV and tuberculosis, little is known about their implementation for NCD-related behavioral interventions within African ED settings^11^ (Tomlinson 2013).

Kenya presents a potentially favorable environment for implementation of mHealth cessation interventions because of widespread mobile phone penetration, increasing digital health infrastructure, and growing interest in technology-supported healthcare delivery. However, successful implementation of innovations across different contexts requires careful adaptation to local workflows, organizational realities, and health system priorities^12^. This study therefore explored key stakeholder perspectives on implementation of the Text2Quit tobacco cessation intervention within a Kenyan ED setting, with particular focus on factors influencing adoption, implementation, sustainability, and scale-up.

## Methods

### Study Design and Setting

This qualitative key informant interview study was nested within a larger type 2 hybrid implementation– effectiveness project designed to assess tobacco use and mHealth cessation readiness among Kenyan ED patients, adapt the Text2Quit program for the local context, and pilot-test the intervention to evaluate its acceptability, feasibility, and preliminary efficacy. Hybrid implementation–effectiveness designs are increasingly used to simultaneously evaluate clinical outcomes and implementation processes^13^.

The study was conducted at Kenyatta National Hospital, a tertiary national teaching and referral hospital located in Nairobi County, Kenya, and the largest public referral hospital in Kenya and East Africa. The hospital serves a large and diverse patient population drawn from Nairobi and referral networks across Kenya and neighboring countries. As a major referral and teaching institution, Kenyatta National Hospital manages a high volume of patients with both acute and chronic conditions, including a substantial burden of non-communicable diseases (NCDs) and tobacco-related illnesses. Prior studies conducted in the hospital’s Emergency Department (ED) have demonstrated a disproportionately high burden of NCDs and associated risk factors among ED populations compared to the general population, highlighting the ED as an important setting for preventive and behavioral health interventions^6,14^.

### Participants and Recruitment

We conducted 12 semi-structured key informant interviews (KIIs) with ED administrators and clinical staff, including departmental heads (n=2), medical doctors (n=5), nurses (n=3), and clinical officers (n=2). A stratified purposeful sampling strategy was used to ensure representation across key professional roles central to ED operations and decision-making.

Sampling was purposive and designed to maximize variation, allowing us to capture a diverse range of perspectives, experiences, and clinical or administrative responsibilities relevant to the adoption, integration, and sustainability of an mHealth tobacco cessation intervention. Although the number of participants was small, this approach prioritized depth and richness of information consistent with qualitative inquiry^15^. Eligible participants were identified from the ED and broader hospital setting by the research team, and recruitment continued until representation was achieved across key professional roles. The final sample size was guided by the principle of data saturation, with recruitment continuing until no new themes emerged from the interviews^16,17^.

### Data Collection and Analysis

Interviews were conducted in English, audio-recorded, and professionally transcribed. Data were analyzed using a combined deductive–inductive approach informed by grounded theory principles, consistent with approaches commonly used in implementation science and qualitative health research^18,19^. The broader hybrid type 2 implementation–effectiveness study was conceptually guided by the RE-AIM framework to examine implementation outcomes related to adoption, implementation, and maintenance of the Text2Quit intervention^20^.

For the qualitative component, select constructs from the Consolidated Framework for Implementation Research (CFIR) informed both the deductive development of the interview guide and the analytic framework used to explore multilevel determinants influencing implementation of the intervention within the ED setting^21^. Deductive coding was guided by relevant CFIR domains and constructs, while inductive analysis allowed unanticipated themes and context-specific insights to emerge from the data and inform interpretation^19^.

Independent coding and analysis were conducted by two trained and experienced qualitative researchers (GJS and LM) with oversight by CN. All transcripts were coded using Dedoose software for systematic data organization, code application, and retrieval. To enhance analytic rigor, coding and interpretation were conducted collaboratively by multiple researchers, with regular consultation meetings and comparison of coded excerpts to discuss coding decisions, emerging themes, and alternative interpretations^22^.

This study was approved by the Kenyatta National Hospital–University of Nairobi Ethics and Research Committee (KNH-UoN ERC). Written informed consent was obtained from all participants prior to participation.

### Researcher Reflexivity and Positionality

The research team included investigators with backgrounds in global health, emergency medicine, implementation science, qualitative research, and non-communicable disease research in Kenyan and international settings. Several members of the team had prior clinical and research experience within Kenyan emergency care settings and longstanding interests in tobacco control and mHealth interventions. These experiences informed interpretation of the findings while also requiring ongoing reflexive consideration of how professional perspectives and prior assumptions could shape data analysis and interpretation.

## Results

### Participant Characteristics

A total of 12 transcripts from Emergency Department (ED) healthcare providers and administrators were analyzed as seen in the demographic table in table 1. Participants were purposively selected from Kenyatta National Hospital, a national referral hospital in Kenya and included individuals directly involved in ED operations and patient care. The sample comprised two departmental heads, five medical doctors, three nurses, and two clinical officers, representing a range of clinical and administrative roles. All participants had direct experience providing care to patients with non-communicable diseases and related risk factors, including tobacco use, and were therefore well positioned to provide insights into the adoption, implementation, and sustainability of a mobile health (mHealth) tobacco cessation intervention within the ED setting.

**Table 1.**
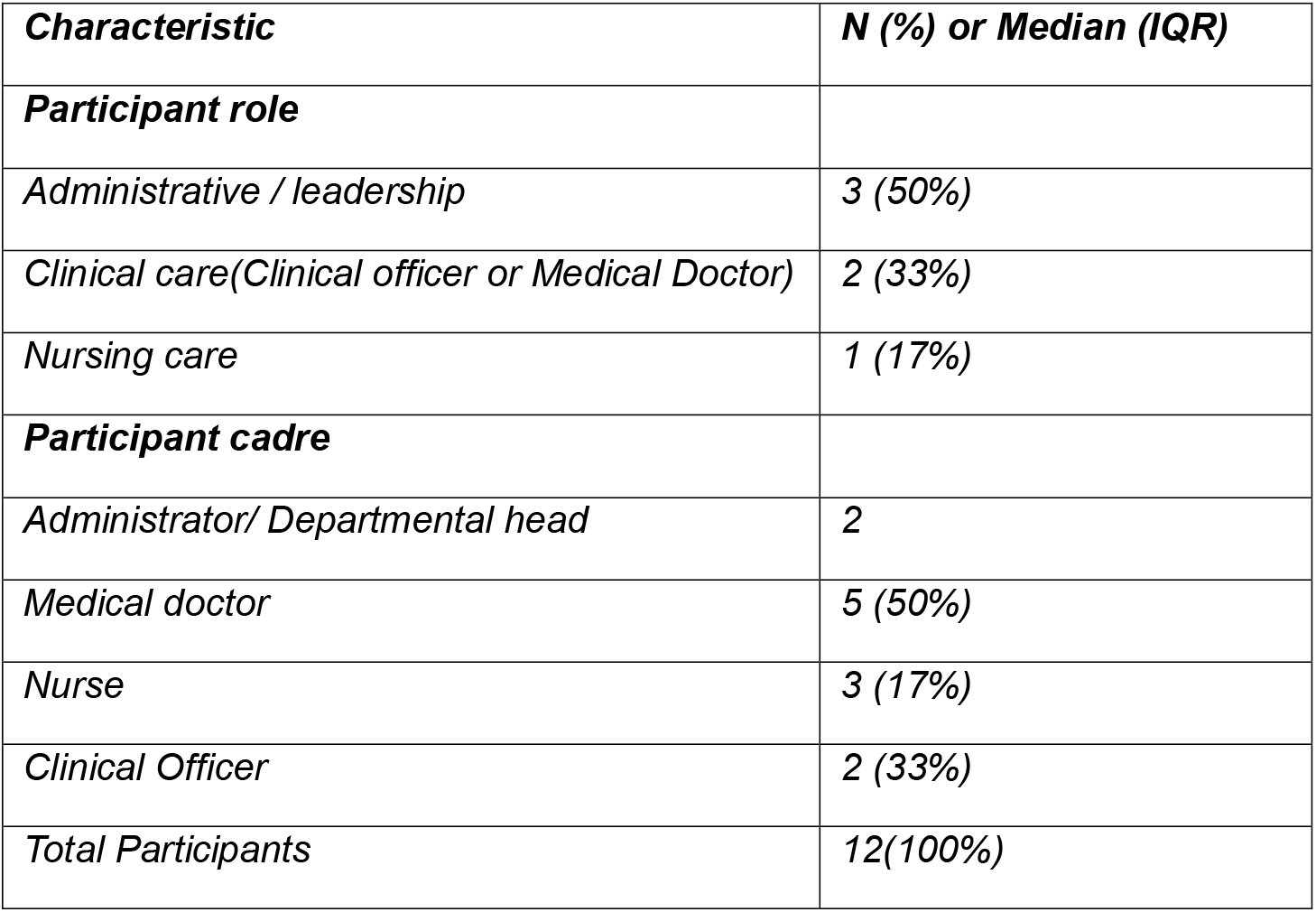
Demographic Table.

### Overview of Findings

Analysis identified several interrelated themes representing multilevel determinants influencing the adoption, implementation, and sustainability of the Text2Quit–Kenya intervention. Guided by a CFIR-informed deductive analytic approach alongside inductive thematic interpretation, findings reflected key domains related to intervention characteristics, inner setting factors, organizational readiness, implementation processes, and patient and provider perceptions influencing integration of the intervention within the ED setting. The broader RE-AIM framework informed interpretation of implementation outcomes related to adoption, implementation, and maintenance within the larger hybrid implementation–effectiveness study.

Themes were identified through iterative analysis of interview data and subsequently mapped to relevant domains of the Consolidated Framework for Implementation Research (CFIR) to further characterize contextual and organizational determinants influencing implementation. Mapping of key findings to CFIR domains is presented in Table 2. Consistent with approaches used in implementation science research, CFIR mapping was used to interpret how multilevel contextual factors shaped perceptions of implementation feasibility, organizational readiness, and sustainability.

**Table 2.** Mapping of Key Findings to CFIR Domains.

| Key Finding | CFIR Domain(s) |
| --- | --- |
| ***Theme 1: Balancing opportunity and feasibility in ED tobacco cessation*** |  |
| Strong perceived need for tobacco cessation services and receptiveness toward the intervention supported adoption | Outer Setting; Inner Setting |
| Simplicity, low complexity, and adaptability of the SMS-based intervention enhanced feasibility and acceptability | Intervention Characteristics |
| Alignment with existing patient education and clinical workflows facilitated integration into routine ED care | Inner Setting |
| Competing clinical priorities, high patient acuity, and pressure to maintain patient flow limited implementation feasibility | Inner Setting |
| Shame, stigma, and reluctance to disclose tobacco use affected patient engagement | Characteristics of Individuals; Outer Setting |
| Resource limitations, including staffing and financial concerns related to SMS delivery, influenced perceptions of feasibility | Inner Setting; Intervention Characteristics |
| ***Theme 2: Strategies for integrating tobacco cessation into routine ED care*** |  |
| Follow-up systems, SMS communication, and outpatient clinic linkage supported continuity of care beyond the ED encounter | Process; Inner Setting |
| Engagement of multiple healthcare cadres and task-sharing approaches facilitated implementation | Process; Inner Setting |
| Patient education and community sensitization were perceived to improve awareness, reach, and engagement | Outer Setting; Characteristics of Individuals |
| Use of implementation champions and reinforcement strategies supported program visibility and accountability | Process |
| ***Theme 3: Conditions required for sustainability and scale-up*** |  |
| Adequate staffing and dedicated funding were viewed as foundational to long-term sustainability | Inner Setting |
| Existing infrastructure, including communication systems, referral pathways, and mobile phone access, supported integration and scalability | Inner Setting; Intervention Characteristics |
| Ongoing staff education, training, and sensitization supported implementation readiness and sustained delivery | Process |
| Institutional policies, leadership engagement, and organizational buy-in enabled long-term integration into routine care | Inner Setting |
| Scalability across facility levels and community settings depended on coordinated systems, standardized processes, and integration into existing care structures | Outer Setting; Process |

Overall, findings highlighted a strong perceived need for tobacco cessation interventions within Kenyan ED settings, alongside recognition that successful implementation would depend heavily on workflow compatibility, organizational support, staffing capacity, and integration into existing care structures. Participants consistently emphasized the importance of minimizing implementation burden through low-complexity intervention design, task-sharing across healthcare cadres, and leveraging existing communication and follow-up systems to support continuity of care and long-term sustainability.

We present findings across three major themes:

1. Balancing opportunity and feasibility in ED tobacco cessation
2. Strategies for integrating tobacco cessation into routine ED care
3. Conditions required for sustainability and scale-up

The code cloud presented in Figure 1 illustrates prominent concepts and implementation determinants influencing the adoption, implementation, and maintenance of the Text2Quit–Kenya intervention.

**Figure 1.**
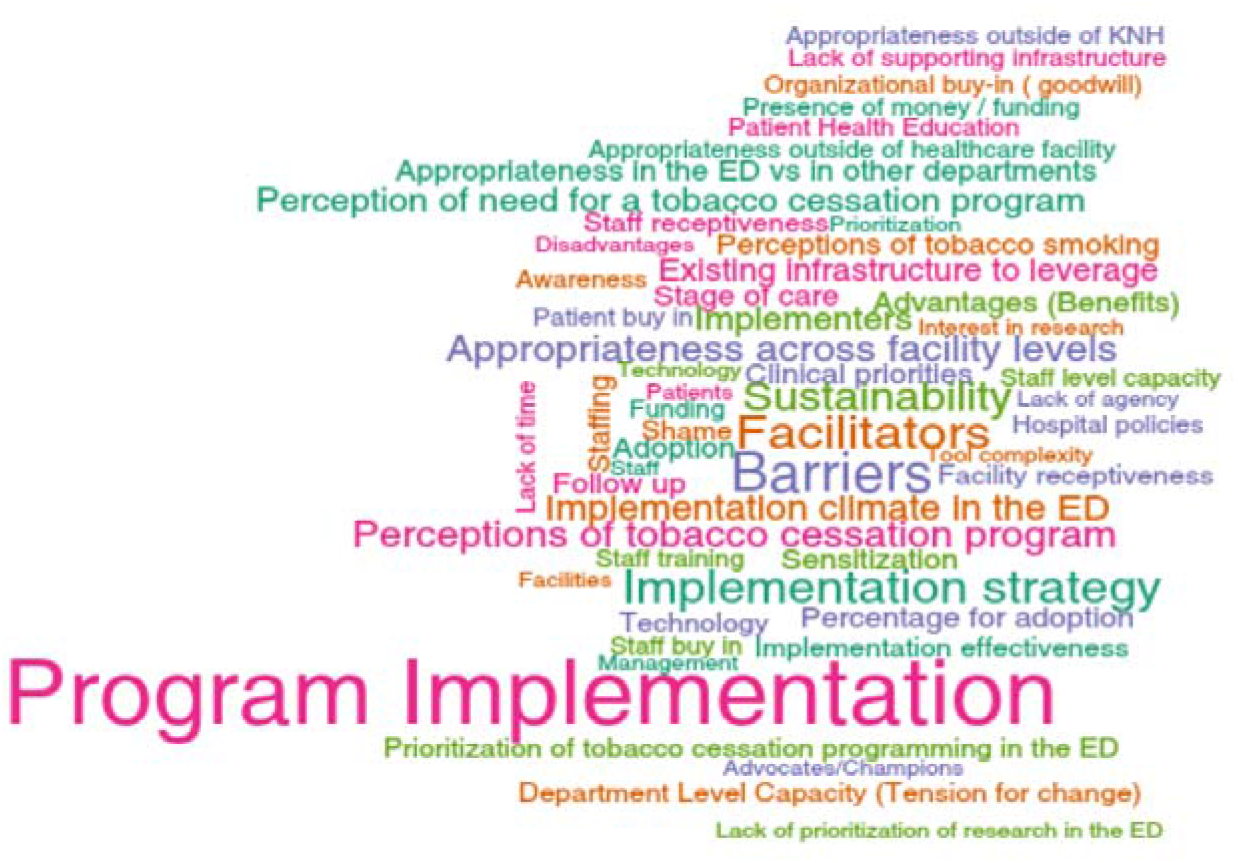
Code / word cloud: concepts and implementation determinants for the Text2Quit–Kenya intervention.

### Theme 1: Balancing opportunity and feasibility in ED Tobacco cessation

Overall, participants expressed strong support for implementation of the mHealth tobacco cessation intervention across Kenyan Emergency Departments (EDs) and broader health system settings. Participants consistently described a substantial unmet need for tobacco cessation support and viewed the intervention as adaptable, scalable, and appropriate across different levels of care within the Kenyan health system. Existing infrastructure, staff receptiveness, and familiarity with mobile phone communication were viewed as important facilitators supporting adoption. Participants also perceived the intervention to align with existing patient education practices and broader preventive health goals within the ED environment.

> *“Ideally, it would be nice if all emergency departments in Kenya utilized this program… we would be able to do a lot of prevention… and enable patients to stop smoking, which will prevent the effects of smoking. We see a lot of patients with cancer of the lungs and throat, and I tend to attribute this to smoking… if we had included this earlier, we would be able to reach a bigger population*.*”*

Despite this overall receptiveness, participants described several contextual and operational barriers that could limit implementation feasibility. Consistent with CFIR domains related to implementation climate and organizational readiness, participants emphasized that the fast-paced nature of emergency care, competing clinical priorities, and pressure to maintain patient flow could reduce opportunities for preventive counseling and enrollment into cessation programs. The acute nature of patient presentations often required providers to prioritize immediate life-threatening conditions over behavioral interventions.

> *“The only thing is at times when patients come in, there are very sick patients in the emergency department, so people tend to focus on what is the main thing that has brought this patient, what can make this patient die or survive. So what I know is the emergency clinicians can be able to forget this aspect, but with practice and as we automate our systems. this has to be something that the doctor has to fill in before they move on to the next stage*.*”*

Financial constraints were also identified as an important barrier to implementation. Participants raised concerns regarding sustainability of SMS delivery costs and broader resource limitations across different levels of the health system, particularly outside tertiary referral settings.

> *“Especially the SMS program, I think the biggest issue would be about monetary. Like who would be funding the EDs in terms of sending the SMSs… majority of level 5, going to level 1 don’t have the resources. If it’s well funded, yes, then people are willing to adopt*.*”*

At the patient level, shame and stigma associated with tobacco use were perceived to potentially limit engagement and disclosure of smoking behavior.

> *“Many people don’t agree that they are smoking… they just come and hide, they don’t tell us that*.*”*

Participants also identified several intervention characteristics that enhanced perceived feasibility and acceptability. Simplicity, low time burden, and ease of integration into existing workflows were consistently viewed as major facilitators of adoption. Interventions requiring minimal provider effort and capable of being embedded into routine intake or discharge processes were perceived as more likely to be accepted within the high-volume ED environment.

> *“If you have a long and complicated tool, people will not have the time or the energy to fill it*.*”*

Participants further emphasized that streamlined and automated systems requiring only essential patient information would improve usability and long-term sustainability.

> *“So, in that aspect I think if you had a tool that is collecting minimal but critical information of the patient’s information… I think that would be a good facilitator*.*”*

Overall, findings from this theme suggest that implementation feasibility was shaped less by resistance to tobacco cessation itself and more by concerns regarding workflow compatibility, operational burden, staffing pressures, and resource demands within the ED setting.

### Theme 2: Strategies for Integrating Tobacco Cessation into Routine ED Care

Participants identified several practical strategies to support integration of the Text2Quit intervention into routine ED care. Across interviews, participants emphasized the importance of embedding the intervention within existing healthcare structures, strengthening patient awareness, and engaging multiple stakeholders to support implementation and continuity of care.

Robust follow-up systems leveraging SMS communication and existing outpatient referral structures were consistently viewed as essential for maintaining patient engagement beyond the initial ED encounter.

Participants described the ED as an important point of entry into the healthcare system and emphasized opportunities to link patients to ongoing care through existing clinics and referral systems.

> *“The KNH emergency department is the first entry point for most patients, so if we capture them there, it is easier. We admit and discharge these patients and have follow-up through chest and other medical clinics… we have the capacity to do that*.*”*

Participants also emphasized the importance of patient education and community sensitization to improve awareness of tobacco-related harms and strengthen engagement with cessation services. Increased sensitization at both facility and community levels was viewed as important for improving reach and program effectiveness.

Stakeholder engagement and inclusion of multiple healthcare cadres emerged as another important implementation strategy. Participants consistently emphasized that responsibility for tobacco cessation activities should not rest solely with physicians, but rather be distributed across nurses, clinical officers, counselors, and other frontline healthcare workers. This approach was viewed as important for reducing provider burden, improving workflow compatibility, and increasing organizational ownership of the intervention.

> *“It doesn’t have to be the doctor or the nurse. Any healthcare worker can be able to sensitize people on the benefits of stopping smoking and the effects of smoking*.*”*

The use of implementation champions and internal advocates was also identified as important for maintaining program visibility, reinforcing adherence, and promoting accountability within the ED setting.

> *“We just need people who will be driving it, maybe posters to remind people. But I think everybody plays a role in it. We obviously need champions to advocate for it in the department*.*”*

Consistent with CFIR process-related domains, these findings suggest that successful implementation would depend not only on the intervention itself, but also on effective stakeholder engagement, communication, workflow integration, and ongoing reinforcement strategies that support normalization of tobacco cessation activities within routine ED care.

### Theme 3: Conditions required for sustainability and scale-up

Participants identified several interconnected organizational and health system factors necessary to support long-term sustainability and scale-up of the Text2Quit intervention. These findings reflected broader considerations related to organizational readiness, leadership engagement, available resources, and infrastructure capacity needed to sustain implementation over time.

Adequate staffing and dedicated funding mechanisms were consistently identified as foundational requirements for sustainability. Participants emphasized that sustained implementation would require sufficient human resources, financial support for SMS delivery, and ongoing investment in implementation activities across different levels of the health system.

> *“Good staffing, everything works, and also money… good money*.*”*

Existing healthcare infrastructure, including outpatient clinics, communication systems, referral networks, and mobile phone access, was viewed as an important facilitator for sustaining and scaling the intervention.

Participants emphasized that leveraging existing systems would support continuity of care and integration into routine healthcare delivery processes.

Participants also identified ongoing education, training, and staff sensitization as essential to maintaining implementation quality and ensuring continued engagement across healthcare cadres. Training was viewed as important not only for building provider confidence, but also for supporting organizational readiness and long-term uptake.

Strong institutional policies, leadership engagement, and organizational buy-in were further identified as key determinants of sustainability. Participants emphasized that embedding the intervention into hospital policies, routine documentation systems, and standardized workflows would strengthen accountability and facilitate long-term integration into ED practice.

> *“Everything is sustainable with good policies… we sustain so many things in the department… all those things are sustainable with good policies, with good staffing, [and] good supervision*.*”*

Participants additionally emphasized that scalability across facility levels and community settings would require coordinated implementation efforts, standardized processes, and alignment with existing healthcare delivery structures. Widespread mobile phone penetration and familiarity with SMS communication were viewed as important enabling factors supporting future expansion beyond tertiary referral facilities.

Collectively, findings from this theme suggest that sustainability of mHealth tobacco cessation interventions in Kenyan ED settings would depend on sustained organizational support, workforce capacity, financing, infrastructure, and integration into routine systems of care.

## Discussion

Implementation of health innovations across different contexts, particularly from high-income to resource-limited settings, requires careful consideration of local health system realities, workflow demands, and contextual determinants that may influence adoption, implementation, and sustainability. Our findings demonstrate that adaptation of the Text2Quit mHealth tobacco cessation intervention for Kenyan Emergency Department (ED) settings was perceived as both feasible and acceptable, particularly when aligned with existing workflows, staffing structures, and communication systems. Participants emphasized that simplicity, low implementation burden, and integration into routine care processes were critical determinants of adoption and sustainability. These findings are consistent with implementation science literature demonstrating that intervention compatibility, complexity, and organizational fit strongly influence implementation outcomes in healthcare settings^12,23^.

Consistent with prior emergency medicine literature, participants recognized the ED as a critical point of contact for high-risk patients with tobacco-related illness and NCD risk factors. Emergency Departments have increasingly been identified as important venues for preventive interventions because they often serve populations with limited engagement in longitudinal healthcare systems^24^. Participants perceived the ED as an opportunity to identify smokers during acute illness encounters and initiate linkage to cessation support. This is particularly important in LMIC settings where access to preventive and behavioral health services may remain limited outside acute care settings.

Our findings also reinforce broader global concerns regarding inequitable access to tobacco cessation support despite implementation of tobacco control policies under the WHO Framework Convention on Tobacco Control^3^. While tobacco control efforts in many high-income countries have contributed to substantial reductions in smoking prevalence, implementation of cessation support services remains inconsistent in many African settings^2^. The WHO MPOWER framework specifically identifies “Offer help to quit tobacco use” as a key evidence-based strategy for reducing tobacco-related morbidity and mortality; however, tobacco dependence treatment services remain among the least implemented tobacco control measures globally, particularly in low-resource settings^25,26^. Participants in this study identified similar structural barriers including staffing shortages, competing clinical priorities, limited financing, and insufficient institutional support as major challenges to implementation. These findings align with prior African tobacco control literature describing inadequate cessation infrastructure, limited provider training, and competing disease priorities as major barriers to tobacco treatment implementation^26,27^.

A particularly important finding from this study was the central role of workflow compatibility and perceived implementation burden in shaping implementation acceptability. Although participants generally perceived the intervention positively, implementation support was conditional on the intervention remaining minimally disruptive to routine ED care. Participants consistently emphasized that interventions perceived as overly time-intensive or operationally burdensome would be unlikely to be sustained in the high-acuity ED environment.

These findings reflect broader implementation science theories emphasizing the importance of normalization, integration into routine practice, and minimization of implementation burden in supporting sustainability of healthcare interventions^23^. The relative simplicity and adaptability of an SMS-based intervention were therefore viewed as major facilitators of implementation feasibility.

Participants identified several practical implementation strategies that could support successful integration of the intervention within routine ED workflows. Follow-up mechanisms leveraging SMS communication and existing outpatient referral systems were viewed as essential for maintaining continuity of care beyond the initial ED encounter. Participants also emphasized the importance of patient sensitization, provider engagement, and organizational awareness-building to improve uptake and normalize tobacco cessation within routine ED practice. These findings align with prior implementation studies demonstrating that stakeholder engagement, implementation champions, and continuous reinforcement strategies improve implementation uptake and sustainability in healthcare settings^28^.

Task-sharing emerged as another important implementation strategy. Participants consistently emphasized that tobacco cessation activities should not rely solely on physicians but instead be distributed across healthcare cadres including nurses, clinical officers, counselors, and other frontline staff. This approach was perceived to reduce provider burden, improve workflow compatibility, and enhance opportunities for patient engagement throughout the continuum of care. Task-sharing approaches have been widely recommended in global health settings as effective strategies for addressing workforce shortages and improving scalability of health interventions in LMICs^29^. Importantly, participants also emphasized the need for staff training and inclusion to support implementation readiness and long-term engagement with the intervention.

The findings further highlight the potential role of low-complexity digital health interventions in expanding access to tobacco cessation support in resource-limited settings. Participants viewed widespread mobile phone access and familiarity with SMS communication as important facilitators of scalability and continuity of care. Unlike smartphone-based applications that may require internet connectivity and higher digital literacy, SMS-based interventions were perceived as feasible across multiple levels of the Kenyan health system. These findings are consistent with broader digital health literature suggesting that simpler mobile interventions may be more scalable and equitable in low-resource contexts^11,30^.

Sustained implementation of the Text2Quit intervention was perceived to depend on several interconnected organizational and system-level factors including adequate funding, staffing, infrastructure, leadership support, and integration into existing workflows. Participants emphasized that long-term sustainability would require embedding the intervention into routine documentation systems, referral pathways, and hospital policies to support accountability and continuity. Organizational buy-in and leadership engagement were viewed as particularly important for ensuring that the intervention became part of routine care rather than a temporary externally funded initiative^12,23^.

Concerns regarding financing of SMS delivery and staffing resources reflected broader sustainability challenges commonly encountered in digital health implementation efforts within LMICs. Prior studies have demonstrated that many digital health interventions in resource-limited settings face challenges transitioning from pilot implementation to sustainable integration within healthcare systems due to limited financing, fragmented infrastructure, and inadequate policy integration^11,30^. Participants in this study similarly emphasized that sustainability would depend not only on availability of technology, but also on continued institutional support, training, and workforce capacity. The widespread availability of mobile phones and existing communication infrastructure were nevertheless viewed as important facilitators that could support future scale-up across multiple levels of care and community settings.

### Gaps and limitations

This study has several limitations. First, interviews were conducted at a single tertiary referral hospital in Kenya, which may limit transferability of findings to other healthcare settings or regions. Second, the study primarily reflects perspectives of healthcare providers and administrators and may not fully capture patient-level experiences and preferences regarding tobacco cessation interventions. Social desirability bias may also have influenced participant responses regarding organizational readiness and receptiveness to implementation. However, the study also has several strengths, including use of implementation science frameworks to guide data collection and analysis, inclusion of multidisciplinary stakeholders, and in-depth qualitative exploration of implementation determinants relevant to ED-based mHealth tobacco cessation interventions in a resource-limited setting.

Overall, the findings suggest that mHealth tobacco cessation interventions may represent a feasible and scalable strategy for expanding access to tobacco dependence treatment within African ED settings. However, successful implementation will likely depend not only on intervention effectiveness, but also on alignment with local workflows, organizational readiness, stakeholder engagement, and availability of sustainable implementation resources. These findings contribute important implementation science insights relevant to integration of preventive behavioral health interventions within emergency care settings in LMICs.

### Implications for D&I Research

Innovation implementation, including innovation transfer between different contexts, requires the inclusion of perspectives of key stakeholders and frontline implementers in co-designing interventions to ensure contextual fit and sustainability^12,21^. Implementing a mobile health tobacco cessation program like Text2Quit in a Kenyan Emergency Department is both feasible and acceptable, but requires targeted strategies such as healthcare provider engagement, program participant follow-up mechanisms, and resource alignment to ensure successful and sustainable integration into the existing healthcare systems.

## Conclusion

Implementing a mobile health tobacco cessation program like Text2Quit in a Kenyan Emergency Department is both feasible and acceptable, but requires targeted strategies such as healthcare provider engagement, program participant follow-up mechanisms, and resource alignment to ensure successful and sustainable integration into the existing healthcare systems.

## Data Availability

All data produced in the present study are available upon reasonable request to the authors

---

**Supplementary Tables 1.** Example coding structure and development of themes.

Supplementary Tables: Example coding structure and development of themes
| Major Theme | Example Supporting Codes |
| --- | --- |
| **Balancing opportunity and feasibility in ED tobacco cessation** | Perceived need for tobacco cessation program |
|  | Facility and staff receptiveness |
|  | Existing infrastructure |
|  | Technology feasibility (SMS communication) |
|  | Tool complexity |
|  | Workflow compatibility |
|  | Competing clinical priorities |
|  | Staffing constraints |
|  | Funding and cost concerns |
|  | Shame and stigma related to tobacco use |
| **Strategies for integrating tobacco cessation into routine ED care** | Follow-up systems and continuity of care |
|  | Patient health education |
|  | Sensitization |
|  | Multidisciplinary implementers |
|  | Task-sharing across healthcare cadres |
|  | Staff training and sensitization |
|  | Champions and implementation advocates |
| **Conditions required for sustainability and scale-up** | Funding and financial sustainability |
|  | Staffing and workforce capacity |
|  | Leadership engagement |
|  | Organizational buy-in and goodwill |
|  | Institutional policies and standardization |
|  | Existing infrastructure to leverage |
|  | Information technology systems |
|  | Continuous education and training |
|  | Scalability across facility levels |

## References

1. World health Organization. WHO - Noncommunicable diseases. 2025.

2. Bilano V, Gilmour S, Moffiet T, D’Espaignet ET, Stevens GA, Commar A, et al. Global trends and projections for tobacco use, 1990-2025: An analysis of smoking indicators from the WHO Comprehensive Information Systems for Tobacco Control. The Lancet. 2015 Mar 14;385(9972):966–76. doi:10.1016/S0140-6736(15)60264-1 PubMed PMID: 25784347.

3. World Health Organization. WHO framework convention on tobacco control. World Health Organization; 2005. 36 p.

4. World Health Organization. WHO report on the global tobacco epidemic, 2008: the MPOWER package. 2008.

5. World Health Organization. WHO global report on trends in prevalence of tobacco use 2000–2024 and projections 2025–2030 [Internet]. 2025. Available from: https://iris.who.int/.

6. Ngaruiya C, Wambua M, Mutua TK, Owambo D, Muchemi M, Rop K, et al. The last frontier for global non-communicable disease action: The emergency department—A cross-sectional study from East Africa. PLoS One. 2021 Apr 1;16(4 April). doi:10.1371/journal.pone.0248709 PubMed PMID: 33798234.

7. Bernstein E, Bernstein J, Feldman J, Fernandez W, Hagan M, Mitchell P, et al. An evidence based alcohol screening, brief intervention and referral to treatment (SBIRT) curriculum for emergency department (ED) providers improves skills and utilization. Subst Abus. 2007 Nov 27;28(4):79–92. doi:10.1300/J465v28n04_01 PubMed PMID: 18077305.

8. Bernstein SL, Bijur P, Cooperman N, Jearld S, Arnsten JH, Moadel A, et al. A randomized trial of a multicomponent cessation strategy for emergency department smokers. Academic Emergency Medicine. 2011;18(6):575–83. doi:10.1111/j.1553-2712.2011.01097.x

9. Free C, Knight RGN R, Robertson SB, Zhou W, Cairns J, Kenward MG, et al. Smoking cessation support delivered via mobile phone text messaging (txt2stop): a single-blind, randomised trial. www.thelancet.com. 2011;378:49–55. doi:10.1016/S0140

10. Abroms LC, Boal AL, Simmens SJ, Mendel JA, Windsor RA. A randomized trial of Text2Quit: A text messaging program for smoking cessation. Am J Prev Med. 2014;47(3):242–50. doi:10.1016/j.amepre.2014.04.010 PubMed PMID: 24913220.

11. Tomlinson M, Rotheram-Borus MJ, Swartz L, Tsai AC. Scaling Up mHealth: Where Is the Evidence? PLoS Med. 2013;10(2). doi:10.1371/journal.pmed.1001382 PubMed PMID: 23424286.

12. Greenhalgh T RGMFBPKO. Diffusion of Innovations in Service Organizations: Systematic Review and Recommendations. 2004.

13. Curran GM, Bauer M, Mittman B, Pyne JM, Stetler C. Effectiveness-implementation hybrid designs: Combining elements of clinical effectiveness and implementation research to enhance public health impact. Med Care. 2012 Mar;50(3):217–26. doi:10.1097/MLR.0b013e3182408812 PubMed PMID: 22310560.

14. Myers JG, Hunold KM, Ekernas K, Wangara A, Maingi A, Mutiso V, et al. Patient characteristics of the Accident and Emergency Department of Kenyatta National Hospital, Nairobi, Kenya: A cross-sectional, prospective analysis. BMJ Open. BMJ Publishing Group; 2017. doi:10.1136/bmjopen-2016-014974 PubMed PMID: 29025826.

15. Patton MQuinn. Qualitative research and evaluation methods. Sage Publications; 2002.

16. Guest G, Bunce A, Johnson L. How Many Interviews Are Enough?: An Experiment with Data Saturation and Variability. Field methods. 2006;18(1):59–82. doi:10.1177/1525822×05279903

17. Saunders B, Sim J, Kingstone T, Baker S, Waterfield J, Bartlam B, et al. Saturation in qualitative research: exploring its conceptualization and operationalization. Qual Quant. 2018 Jul 1;52(4):1893–907. doi:10.1007/s11135-017-0574-8 PubMed PMID: 29937585.

18. Fereday J, Adelaide N, Australia S, Eimear Muir-Cochrane A. Demonstrating Rigor Using Thematic Analysis: A Hybrid Approach of Inductive and Deductive Coding and Theme Development. 2006.

19. Charmaz K. Grounded Theory.

20. Glasgow RE, Vogt TM, Boles SM, Glasgow E. Evaluating the Public Health Impact of Health Promotion Interventions: The RE-AIM Framework [Internet]. Vol. 89. 1999. Available from: www.ori.

21. Damschroder LJ, Reardon CM, Widerquist MAO, Lowery J. The updated Consolidated Framework for Implementation Research based on user feedback. Implementation Science. 2022 Dec 1;17(1). doi:10.1186/s13012-022-01245-0 PubMed PMID: 36309746.

22. Bradley EH, Curry LA, Devers KJ. Qualitative data analysis for health services research: Developing taxonomy, themes, and theory. Health Serv Res. 2007 Aug;42(4):1758–72. doi:10.1111/j.1475-6773.2006.00684.x PubMed PMID: 17286625.

23. May C, Finch T. Implementing, embedding, and integrating practices: An outline of normalization process theory. Sociology. 2009;43(3):535–54. doi:10.1177/0038038509103208

24. Boudreaux ED, Camargo CA, Arias SA, Sullivan AF, Allen MH, Goldstein AB, et al. Improving Suicide Risk Screening and Detection in the Emergency Department. Am J Prev Med. 2016 Apr 1;50(4):445–53. doi:10.1016/j.amepre.2015.09.029 PubMed PMID: 26654691.

25. World Health Organization (WHO). WHO best buys for the NCDs [Internet]. 2023 [cited 2024 Nov 16]. Available from: https://www.who.int/news/item/26-05-2023-more-ways--to-save-more-lives--for-less-moneyworld-health-assembly-adopts-more-best-buys--to-tackle-noncommunicable-diseases

26. World Health Organization. WHO report on the global tobacco epidemic, 2023_protect people from tobacco smoke. 2023.

27. Drope Jeffrey, Schluger NW. The tobacco atlas. American Cancer Society□; Vital Strategies; 2018. 56 p.

28. Aarons GA, Hurlburt M, Horwitz SMC. Advancing a conceptual model of evidence-based practice implementation in public service sectors. Administration and Policy in Mental Health and Mental Health Services Research. 2011 Jan;38(1):4–23. doi:10.1007/s10488-0100327-7 PubMed PMID: 21197565.

29. Joshi R, Alim M, Kengne AP, Jan S, Maulik PK, Peiris D, et al. Task shifting for non-communicable disease management in low and middle income countries - A systematic review. PLoS ONE. Public Library of Science; 2014. doi:10.1371/journal.pone.0103754 PubMed PMID: 25121789.

30. Labrique AB, Vasudevan L, Kochi E, Fabricant R, Mehl G. mHealth innovations as health system strengthening tools: 12 common applications and a visual framework. Global Health: Science and Practice [Internet]. 2013. Available from: www.ghspjournal.org

